# T2-weighted T1 mapping and automated segmentation of CSF: Assessment of solute gradients in the healthy brain

**DOI:** 10.1101/2025.07.19.25331835

**Authors:** Tryggve Holck Storås, Sofie Lysholm Lian, Ingrid Mossige, Jørgen Riseth, Siri Fløgstad Svensson, Grethe Løvland, Geir Ringstad, Kent-André Mardal, Kyrre Eeg Emblem, Kaja Nordengen

**Author notes:** **Corresponding authors:** PhD Tryggve Holck Storås, Department of Physics and Computational Radiology, Oslo University Hospital, Gaustad Oslo, Norway PO 4950 Nydalen, 0424 Oslo, MD PhD Kaja Nordengen, Department of Neurology, Oslo University Hospital Rikshospitalet, PO 4950 Nydalen, 0424 Oslo.

## Abstract

**BACKGROUND:** Cerebrospinal fluid (CSF) serves as a vehicle for nutrient delivery and waste clearance. The T1 relaxation rate R1 can be used to measure concentration of intrinsic solutes and extrinsic contrast agents.

**PURPOSE:** To implement a method for R1 mapping and segmentation of CSF and to use this method to explore how R1 of CSF vary with CSF protein content and Gadobutrol after intrathecal administration.

**STUDY TYPE:** Prospective cohort study, complemented by phantom analysis.

**POPULATION:** Ten healthy control subjects (mean age 65.5±4.4 years, range 57-72 years; five males and five females) and a protein-gradient phantom for study validation.

**FIELDSTRENGTH/SEQUENCE:** 3T Philips Ingenia scanner; 3D T2W mixed inversion recovery spin-echo (T2W Mixed IRSE) sequence and 3D T1W turbo field echo (3D T1W-TFE).

**ASSESSMENT:** R1 maps were calculated by combining inversion recovery and spin echo data. An automated segmentation method derived from FreeSurfer employed spin-echo data for CSF segmentation and T1W-TFE for anatomical reference. Through lumbar puncture, CSF was harvested for total protein measurements, and 0.25mmol Gadobutrol were injected intrathecally. Post contrast assessments were performed at 6h, 24h, 48h and 72h.

**STATISTICAL TESTS:** One-way ANOVA, followed by a post-hoc Tukey HSD test, and simple linear regression analysis; significance level of 0.05.

**RESULTS:** Intrinsic R1 values in the subarachnoid space were significantly higher than that in ventricles (p < 0.001) and correlated with lumbar protein concentration (p < 0.05). Peak Gadobutrol concentrations were 80 ± 66 µM in ventricles, 146 ± 70 µM in cerebellar SAS and 131 ± 72 µM in cerebral SAS. Corresponding concentrations were 5.2 ± 4.3 µM, 13.0 ± 6.9 µM and 30 ± 15 µM at 72 hours after administration.

**DATA CONCLUSION:** Intrinsic R1 of CSF in subarachnoid space correlated with protein content. Intracranial CSF enrichment after intrathecal administration of Gadobutrol showed a large variation among healthy volunteers.

## Introduction

Cerebrospinal fluid (CSF) plays a crucial role in brain health, acting as a medium for delivery of nutrients and drugs as well as waste clearance, thereby maintaining the homeostasis of the central nervous system by removing toxic by-products of metabolism [1]. Aberrations in CSF dynamics are believed to contribute to neurological conditions, including neurodegenerative disorders [2].

Recent studies have revealed that CSF interacts with interstitial fluid through a brain-wide network of perivascular spaces known as the ‘glymphatic’ system, which present a potential pathway for the clearance of proteins and metabolic waste [3, 4]. A study sampling CSF in felines found gradients of solutes and protein content between the ventricular system and the subarachnoid space (SAS), with three times higher total protein concentration in SAS as compared to VS [5]. Similar patterns are seen in humans [6, 7].

As proteins modify T1 relaxation rates [8], spatial distribution of proteins in CSF might be assessed by MRI provided sufficiently precise T1-mapping, and proper accounting for other sources of T1 relaxation is obtained. Such *In vivo* assessment of protein content in intracranial CSF, and possibly also within the spinal CSF, may provide a better understanding of CSF physiology, including inter-individual variations. It may non-invasively add to diagnostic workups to characterize altered protein content in CSF.

CSF-mediated clearance of endogenous solutes from the brain may also be assessed using an extrinsic MRI contrast agent as CSF tracer, serving as surrogate marker of larger metabolites such as amyloid-beta and tau. Intrathecal contrast enhanced MRI, also referred to as glymphatic MRI (gMRI), have shown that CSF-tracer distribution and clearance within the ventricles, cisterns, subarachnoid space and parenchyma vary significantly between diseases and subjects [9, 10]. The CSF tracer exchanges substantially with the extra-vascular compartment in the brain, and based on T1-weighted images, which are not quantitative per se, it was estimated that on average around 20% of the injected dose was within the parenchyma at peak enrichment [11]. The results aligned with estimates based on quantitative T1-maps in a single subject [12].

The gMRI method’s ability to assess enhancement and clearance properties of parenchyma relies on distribution of the contrast agent from the lumbar injection point to the CSF traversing the investigated tissue. For quantification, the local concentration of contrast agent in CSF must be established using T1 measurements. Such measurements pose a challenge as T1 of CSF is much longer than that of brain tissue. Simultaneous measurements of tissue T1 and CSF T1 would require very long scan times, and at the tissue –CSF interface partial volume effects would be a challenge to standard T1-mapping methods. T2 is also significantly longer in CSF than in tissue. The long T2 of CSF may be exploited in two ways. For one, partial volume effects may be minimized by sampling at long echo times, where parenchymal signal has largely decayed, and secondly, long echo time trains may be used with little T2-blurring, allowing high resolution imaging within a reasonable scan-time.

In this work, we apply a compressed SENSE –accelerated, heavily T2-weighted mixed inversion recovery and spin-echo (T2W-mixed IRSE) 3D-acquisition to measure T1 in CSF with high resolution and full cranial coverage. We implement an automatic segmentation tool for CSF analysis. We also perform a phantom study to establish the relaxivity of albumin at 3T and compare this to observed variations of intrinsic R1 in CSF correlated to *in vitro* protein measurements of lumbar CSF. The aim was to explore variations in intrinsic T1 of CSF and its relation to protein content, and to characterize the distribution of contrast agent after intrathecal administration in healthy volunteers.

## Materials and Methods

### Ethics

The study obtained approval from the Regional Ethics Committee (REC #282297) and the Hospital Authority (Data Protection approval #21/19051). Written informed consent was obtained from all participants, ensuring they understood the study’s purpose, procedures, and their rights, including the right to withdraw at any time.

### Cohort

A total of ten healthy control subjects were included, with a mean age of 65.5 years (range 57-72 years, SD = 4.36), balanced by sex (five females and five males). Inclusion criteria required participants to be healthy individuals aged between 50 and 75 years. Exclusion criteria encompassed reduced kidney function, a history of severe allergies, severe neurological disease, antithrombotic treatment, and claustrophobia.

Participants were primarily recruited through the Norwegian Parkinson Foundation as part of the GRIP study on glymphatic clearance via public presentations and webinars, and involves mostly spouses of Parkinson’s patients. Baseline MRI scans preceded the intrathecal administration of Gadobutrol (0.25 mmol). After intrathecal injection of Gadobutrol, the participants were instructed to stay in bed until 3 p.m. Subsequent MRI exams were conducted at approximately 6, 24, 48, and 72 hours post-administration. CSF was sampled prior to the contrast injection, analyzed for leukocytes, protein, and glucose levels, and stored in a biobank for future research.

### Artificial cerebrospinal fluid

To study the effect of proteins in CSF, we established a semi-logarithmic concentration gradient of human serum albumin (HSA, Sigma-Aldrich, USA, CAS 70024-90-7) within modified artificial CSF (aCSF), tailored to encompass the physiological protein concentration range found in human CSF. HSA is the predominant protein in CSF accounting for over half of the total protein content, which typically varies from 200 to more than 600 mg/L[13]. Our five-step gradient was designed to extend both below and above this physiological window, with concentrations set at 0, 100, 500, 1000, and 5000 mg/L. We prepared these concentration points in duplicate, excluding the mid concentration of 500 mg/L. The composition of our aCSF was adapted from the formulation by Steffensen et al. (2023) [14], with the following modifications to more accurately simulate human CSF: we decreased the glucose concentration from 10 to 3 mM to better reflect the mean glucose levels found in human CSF [15] and increased the concentration of N-2-hydroxyethylpiperazine-N’-2-ethanesulfonic acid (HEPES) from 17 to 20 mM to enhance pH stability [16]. The final HEPES-buffered aCSF solution contained the following components (in mM): 120 NaCl (Sigma-Aldrich, USA, S3014), 2.5 KCl (Sigma-Aldrich, USA, P-9541), 2.5 CaCl2 (Sigma-Aldrich, USA, C-7902), 1.3 MgSO4 (Sigma-Aldrich, USA, M5921), 1 NaH2PO4 (Merck, Germany, A689746633), 3 glucose, and 20 Na-HEPES (Sigma-Aldrich, USA, H-7006), resulting in osmolarity of 300 mOsm/L.

### MRI Sequence

Imaging was done at a 3T Philips Ingenia scanner (Philips Healthcare, Best, The Netherlands) with a 32-channel head coil. This work reports results based on two acquisitions. A T2W-mixed IRSE sequence as shown in the sequence diagram in Figure 1 was used for T1 mapping and segmentation of the intracranial CSF spaces and a 3D T1-weighted TFE was used for anatomical reference. Scan parameters are listed in Table 1. Both acquisitions were sagittal and covered the full anatomy. Nonselective excitation and refocusing pulses were applied. Repetition time of the IR sequence was set so that time for free relaxation was the same between the SE- and IR-sequence. Scan duration for the T2W-mixed IRSE was 7:44 min.

**Table 1.** Scan parameters for the sequences used. NA: Not applicable.

| Scan parameter | 3D IR-TFE | 3D T2W-mixed IRSE |
| --- | --- | --- |
| Repetition time (inversion recovery/spin-echo) | 5.2 ms | 11000 ms/8350 ms |
| Shot interval | 3000 ms | NA |
| Inversion time | 853 ms | 2650 ms |
| Echo time | 2.3 ms | 700 ms |
| Flip angle/Refocusing angle | 8/NA degrees | 90/180 degrees |
| Field of view | 256 x 256 x 184 mm | 224 x 224 x 180 mm |
| Measured voxel size | 1 mm, isotropic | 1 mm, isotropic |
| Reconstructed voxel size | 0.5 mm, isotropic | 0.5 mm, isotropic |
| Bandwidth | 394 Hz/pixel | 286 Hz/pixel |
| Compressed-sense acceleration | 2 | 9 |
| Total scan time | 4:10 min | 7:44 min |
| TFE-factor | 232 | NA |
| Turbo spin echo factor | NA | 166 |
| Dummy echoes | Default | 20 |

**FIGURE 1.**
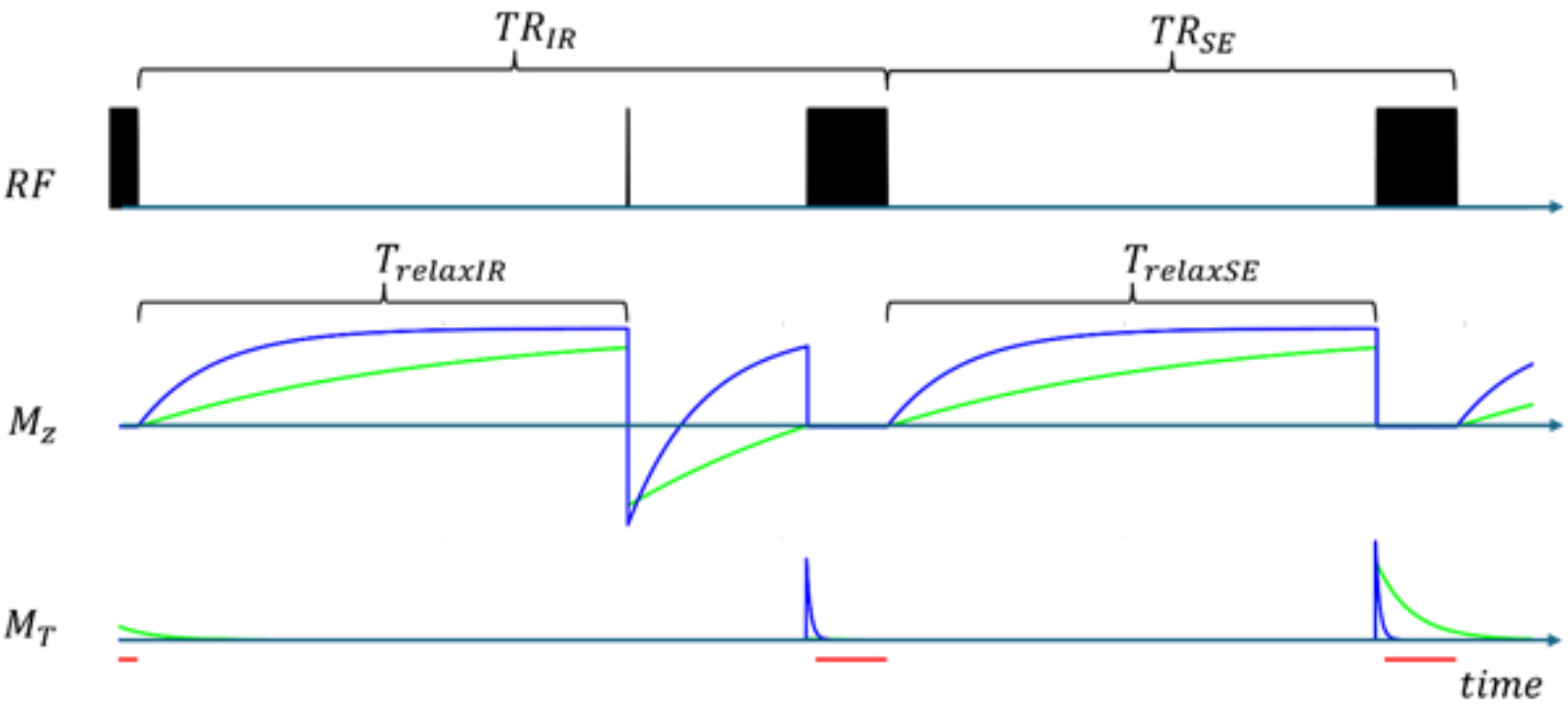
Diagram showing a full cycle of the T2W-mixed IRSE sequence. The top row shows applied radiofrequency (RF) pulses. Durations TR_IR_ and TR_SE_ of the respective IR and SE blocks are set so that time for free relaxation T_relaxIR_ and T_relaxSE_ are equal. Bloch simulations of CSF (green line) and parenchyma (blue line) show how longitudinal magnetization (second row) of all tissues is effectively nulled by the TSE readout. Selection of the CSF signal only is done by limiting the readout (red line) to later echoes where transverse magnetization (third row) of parenchyma is largely decayed.

In addition to the above, the baseline protocol included standard clinical sequences and sequences aimed at gMRI [9]: FLAIR, SWI, PD-Black-blood, T2W, Look-locker and DTI amounting to a total scan duration of 59 minutes, The later sessions used a subset of these scans amounting to a total scan time of 19 minutes per session.

### Post processing

The signal intensities S_SE_ and S_IR_ of the SE- and IR images, respectively, are assumed to follow the signal equations

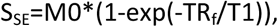

and

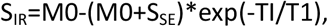

where M0 is the equilibrium magnetization, TI is the inversion time and TR_f_ is the time for free relaxation defined by the repetition time of the spin-echo acquisition minus the echo train duration. The ratio between the signal equations, S_IR/_S_SE_, is a monotonically decreasing function of the T1 time which may be inverted to generate T1 relaxation maps. A lookup table for the expected ratio was created for integer values of T1 from 100 ms to 10000 ms. The ratio between the measured signal intensities was computed for each image voxel and consecutively mapped to the T1 time corresponding to the nearest value from the lookup table. We note that S_SE_ was read from modulus images while S_IR_ was read from phase-corrected real images to preserve polarity.

As T1 relaxation rate R1 = 1/T1 vary in a linear fashion with concentration of solutes, T1 maps were converted to R1-maps by inversion and R1 was used throughout the analysis. Each of the R1 maps for a given subject were co-registered to the corresponding pre-contrast T1-weighted image, using the software *greedy* (https://github.com/pyushkevich/greedy co-registration in ITK-SNAP [17]).

### Segmenting the subarachnoid space

The CSF mask was generated by applying a thresholding algorithm to the pre-contrast S_SE_-image. We used Li’s thresholding algorithm [18, 19] as implemented in Scikit-image [20]. After initial thresholding, we removed all but the largest connected region of the mask. Two voxels were considered connected if one can be reached from the other with a maximum of 2 orthogonal steps.

For each of the study’s subjects, the pre-contrast T1-weighted TFE scans were processed using the recon-all pipeline in FreeSurfer[14] to generate brain segmentations. We relied on the FreeSurfer segmentation aparc+aseg.mgz which combines the cortical parcellations of the Desikan-Killiany atlas [[21],[22]] and the FreeSurfer subcortical segmentation [[23]]. After we removed several labels from the original segmentation, which we deemed irrelevant for the SAS segmentation, the brain segmentations were extrapolated to the voxels of the CSF-mask to segment the SAS [18-20]. We assigned the label of the nearest labelled voxel in the brain segmentation to each voxel in the SAS mask, using the NearestNDInterpolator implemented by Scipy (version 1.15) [24]. Finally, we grouped different SAS segments into either ventricles, cerebral SAS or cerebellar SAS. The code, allowing for customization, is available at (https://github.com/jorgenriseth/grip-mixed.git)

### Estimating amount of Gadobutrol in CSF

We estimated concentration of Gadobutrol from the formula:

[Gadobutrol]=(R1(t)-R1(0))/r1,

where R1(t) is R1 measured at time t, R1(0) is the intrinsic R1 as measured in the baseline examination and r1=4 L/(mmol ·s) is the assumed relaxivity of Gadobutrol in CSF. The total amount of Gadobutrol present in cranial CSF was estimated by multiplying the mean estimated concentration by the volume of the CSF segmentations.

### Reducing high intensity rim effects for R1 estimates

A rim of high R1 was seen at the interface between CSF and gray matter. To investigate the effect of this phenomenon we tested repeated erosion operations on the segmentations while calculating mean R1. Based on this we chose to remove two voxel layers from the segmentations described above before R1 was extracted.

### Statistical methods

For each participant, the mean R1 in three CSF regions (cerebral SAS, cerebellar SAS and ventricles) was calculated excluding the 30 % highest and lowest values. Excluding the 30% highest and lowest values ensure that the mean is more robust within the eroded volume.

Statistical analyses and graphical visualizations, including boxplots, were generated using R version 4.3.2 [25] in the RStudio version 2023.06 [26]. A one-way ANOVA, followed by a post-hoc Tukey HSD test was used to compare R1-values in different CSF regions. The relationship between R1-values in different CSF compartments and lumbar protein concentration was analyzed using simple linear regression. For each CSF region, outliers in the regression analysis were identified using Cook’s distance. Samples with a Cook’s distance greater than 4 / (N ™ k ™ 1), where *N* is the total number of samples and *k* is the number of predictors, were removed. In cases with multiple outliers, the most extreme outlier was removed first, and the detection procedure was repeated until no further outliers were identified. Simple linear regression analysis was also used to compare HSA concentrations and R1-values in artificial CSF. A statistical significance level of 0.05 was used.

A one-sided paired T-test (p < 0.05) was used to test for Gadobutrol enhancement at 72 hours.

Data points and lines were color coded by volunteers to facilitate visual correlation between intrinsic R1, protein data and contrast-enhanced R1 data.

## Results

Intrinsic R1 of CSF was higher in SAS compared to ventricles, and in cerebral SAS compared to cerebral SAS shown for all ten participants in Figure 2A together with an example of the automatically segmented compartments in Figure 2B. Mean estimated cerebral CSF volume was 427 ± 92 (range 256 to 554) ml, of which cerebral SAS contributed 329 ± 67 (range 193 to 419) ml and ventricles 44 ± 19 (range 25 to 90) ml. Volumes and R1s of eroded segmentations are shown in Supplements Figure 1. R1 estimates decreased after the two first voxel erosions, before stabilizing.

**FIGURE 2.**
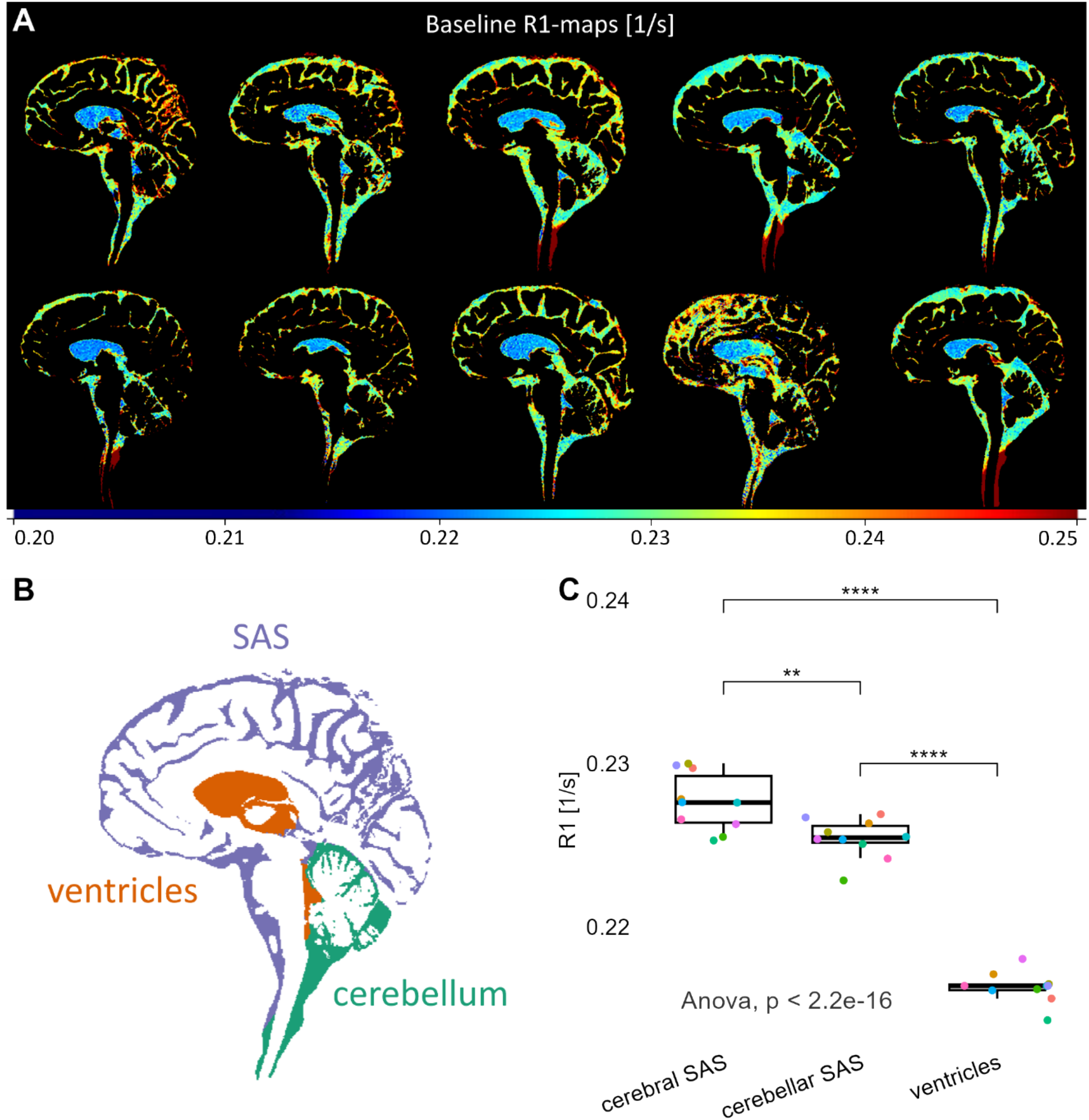
Sagittal R1-maps of all ten volunteers shown with a common compressed color-scale to facilitate comparison and bring forth the subtle but still robust compartmental variation (A). Automatically defined segmentations collected into three main regions comprising mainly CSF in SAS surrounding the cerebrum (purple), the cerebellum (green), and the ventricles (orange) are shown for one volunteer in (B). Estimated mean R1 per volunteer in these segmentations are shown by colored dots (C) and distributions over all volunteers are summarized by boxplots. Stars signify statistically significant (**: p<0.01, ****: p<0.0001) differences by post-hoc Tukey HSD pairwise comparisons as delineated by the brackets.

After erosion of outer two layers R1 of ventricular CSF was 0.216 ± 0.001 s^−1^ which was significantly lower than that of CSF in SAS surrounding the cerebellum 0.225 ± 0.001 (p <0.0001) and cerebrum 0.228 ± 0.002(p < 0.0001), as shown in boxplot in Figure 2C.

Results from regression analysis on HSA in artificial CSF are shown in Figure 3 (A - B). Relaxivity r1 of HSA in artificial CSF was estimated to 0.0019 ± 0.001 L(sg)^−1^ at 37°C, showing a significant correlation between estimated R1 from T2W-mixed IRSE and HSA concentrations in aCSF.

**FIGURE 3.**
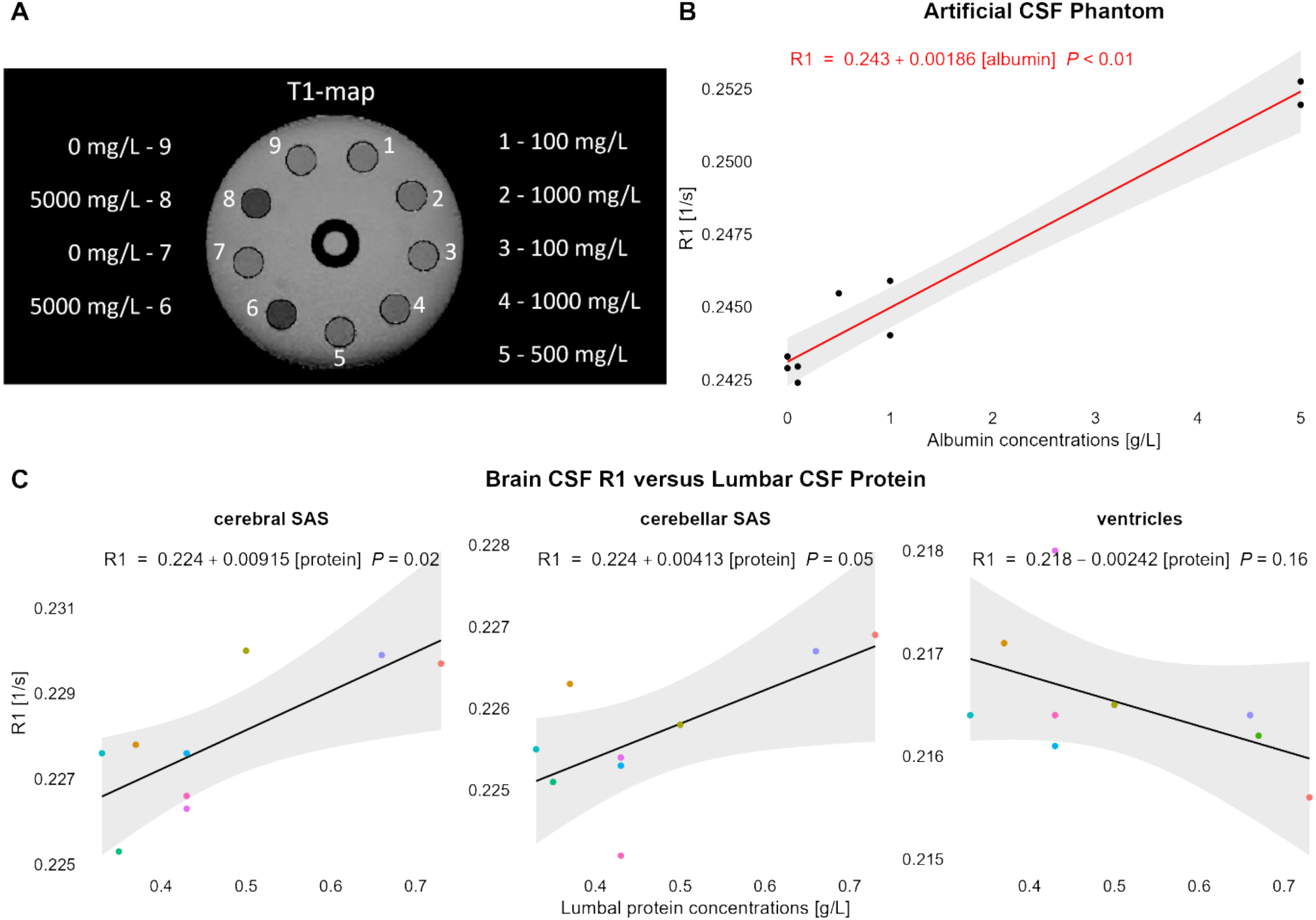
T1 map of phantom containing artificial CSF with a range of human serum albumin concentrations (A). Measured R1 at 37°C plotted against corresponding albumin concentrations was used to estimate the relaxivity r1 of albumin by linear regression (B). Paired samples show a noticeable scatter. Red line shows regression line and gray area shows 95% confidence intervals. *In vivo* measured R1 in CSF surrounding cerebrum, cerebellum and in the ventricles before contrast injection are plotted against corresponding protein concentrations in lumbar CSF (C). Black lines show regression line. Gray areas show 95% confidence intervals. Each volunteer is shown with a unique color which corresponds to Figure 2 and 4.

Regression results relating estimated R1 in eroded segments to CSF protein concentrations as measured by lumbar CSF samplings are shown in Figure 3 C and Table 2. One outlier was detected and excluded from analysis in each region. Correlations were significant for both SAS-regions. The estimated slope was highest in cerebral SAS r = 0.00915 ± 0.0030 L(sg)^−1^. In cerebellar SAS r= 0.0041 ± 0.0017L(sg)^−1^ was still higher than r1 estimated for HSA in aCSF. A negative slope r =- 0.0024 ± 0.0015 L(sg)^−1^ was seen in ventricular CSF. This did not reach statistical significance.

**Table 2.** Simple linear regression analysis of R1-values in different CSF compartments versus lumbar protein concentration.

|  | Intercept<br>[1/s] | SE<br>[1/s] | Estimated<br>r1 [L/sg] | SE [L/sg] | t value | P(>\|t\|) |
| --- | --- | --- | --- | --- | --- | --- |
| Cerebral SAS | 0.2236 | 0.0015 | 0.0091 | 0.0031 | 3.00 | 0.02 |
| Cerebellum SAS | 0.2237 | 0.0008 | 0.0041 | 0.0017 | 2.43 | 0.05 |
| Ventricles | 0.2177 | 0.0007 | -0.0024 | 0.0015 | -1.59 | 0.16 |

Time evolution of R1 after intrathecal administration of Gadobutrol is shown in Figure 4. There was substantial inter-individual variance in tracer distribution. Seven out of ten volunteers showed enhancement in the ventricles at 6 hours post contrast administration. Peak delta R1 in ventricles was 0.32 ± 0.27 (range 0.00 to 0.77) s^−1^ at 6 hours. In cerebellar SAS peak delta R1 was 0.58 ± 0.28 (range 0.15 to 1.00) s^−1^ and in cerebral SAS 0.52 ± 0.28 (range 0.12 to 1.06) s^−1^, both at 24 hours. Assuming a relaxivity of Gadobutrol of 4 (mM s)^−1^, this translates to mean peak Gadobutrol concentrations of 80 ± 66 µM in ventricles, 146 ± 70 µM in cerebellar SAS and 131 ± 72 µM in cerebral SAS.

**FIGURE 4.**
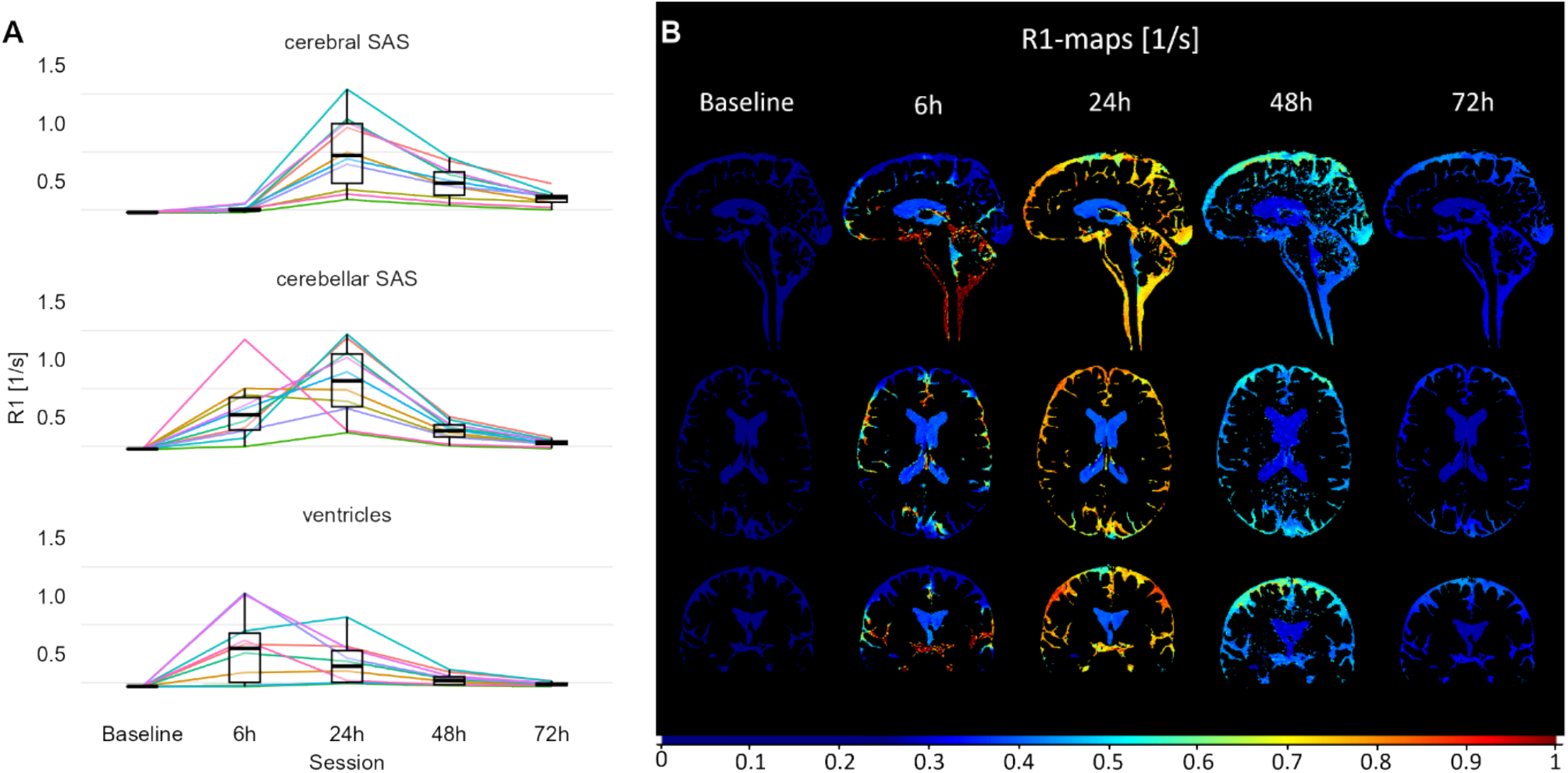
Time evolutions of R1 over all volunteers are shown by segmented regions (a). Colored lines trace individual participants. There was substantial inter-individual variance in tracer distribution among these healthy controls. Seven out of ten volunteers showed significant enhancement in the ventricles at 6 hours post contrast administration. SAS surrounding the cerebellum showed some enhancement at 6 hours but peaked at 24 hours while SAS surrounding cerebrum was largely unaffected before it peaked at 24 hours. After 24 hours R1 of all three regions gradually decreased. At 72 hours R1 in ventricles was close to its baseline value but there was still significantly increased R1 in all regions. Temporal and spatial mapping is shown for one volunteer in (B). The color scale span R1 from 0 to 1 s^−1^.

After 24 hours, R1 of all three regions gradually decreased. At 72 hours, mean enhancement was still significant in all segments. Mean delta R1 was 0.021 ± 0.017 s^−1^ in ventricles, 0.052 ± 0.028 s^−1^ in cerebellar SAS and 0.121 ± 0.064 s^−1^ in cerebral SAS which translates to 5.2 ± 4.3 µM, 13.0 ± 6.9 µM and 30 ± 15 µM of Gadobutrol respectively.

At 24 hours post injection mean estimated total amount of Gadobutrol within cranial CSF was 15 ± 7 (range 4 to 24) % of administered dose.

## Discussion

In this study we applied a T2W-mixed IRSE MRI sequence to measure intrinsic and gadolinium-enhanced T1 relaxation in human CSF. We used a long echo-time to minimize partial volume effects and to allow T2-based segmentation and T1-mapping to be done on the same data. A persistent compartmental pattern was evident in all ten volunteers showing a lower intrinsic R1 of CSF in ventricles than in SAS. We obtained lumbar protein concentrations for correlation with intrinsic R1 and we performed a phantom study to assess the T1-relaxivity of albumin, the most abundant protein in CSF. We found significant correlations between estimated R1 in both cerebral and cerebellar SAS and protein concentration in lumbar CSF, in addition to a significant correlation between the protein gradient in aCSF and estimated R1 in the phantom study. After administration of intrathecal Gadobutrol we found a large variation in enhancement among the healthy volunteers. The availability of a heavily T2-weighted 3D image volume allowed for robust segmentation of CSF, and we developed an automated segmentation of CSF volumes and sub-volumes freely available at GitHub.

Compartmental differences have been reported for both T1 and T2 of CSF [27, 28]. The source of this effect may be multi-faceted. The subarachnoid trabeculations may play a role as they increase the tissue surface CSF is exposed to in SAS, possibly facilitating relaxation by water adsorption [29] or exchange also outside the brain-CSF border.

Using ultra-long-TE arterial spin labeling, Petitclerc et al. [30] estimated a water exchange between blood and CSF in the SAS, occurring at a rate of approximately one per minute. Later, Jiang et al. [31] measured a difference in T1 of CSF in SAS and the frontal horn of the lateral ventricles. They assumed negligible exchange between parenchyma and CSF in the frontal horns and attributed the lower T1 in SAS to water exchange between parenchyma and SAS. Using the Bloch equation including exchange, they found that the T1 difference was adequately explained by water exchange rates in the range 1 to 2 per minute, compatible with the results of Petitclerc. The T1 found by Jiang was equal within statistical errors to the T1 we found before eroding the segmentations.

We observed a rim of high R1 at the interface between CSF and gray matter. This may be caused by residual gray matter signal at the beginning of sampling. To reduce this effect, we used 20 dummy echoes as a compromise between artifact level and scan efficiency. The remaining effect was handled by eroding the segmentations before statistics were calculated.

Water exchange between CSF and gray matter could presumably also cause a high R1 rim as we observe. Assessment of this with the applied sequence would require detailed accounting for tissue point spread functions and was outside the scope of our study.

Solutes in CSF may also contribute to the observed compartmental differences. R1 of CSF surrounding the cerebellum was intermediate, consistent with mixing of fresh CSF from ventricles with low R1 and CSF circulating in the cerebral SAS with higher R1. This apparent dilution effect indicates that CSF solutes also have a considerable impact on intrinsic R1 of CSF.

Ventricular CSF is known to have a low protein concentration compared to lumbar CSF [7]. We found no studies reporting on protein concentrations in cranial SAS in humans. This lack of data may be due to the invasiveness of sampling and the common assumption that mixing of CSF allows presumably representative measurements to be performed in the more available lumbar CSF.

We measured the relaxivity r1 of HSA in aCSF. This was lower than previously reported by Yilmaz et al. for serum proteins at 1T [32]. The albumin thumbling frequency of about 50 MHz[33] is close to optimal for inducing relaxation at 1T and a reduced relaxivity is expected as the Larmour frequency is increased.

Comparing correlations between R1 in SAS and ventricles with lumbar protein concentrations we found regression slopes to be steeper than predicted by the phantom study and that the slope of cerebral SAS was about twice that of the cerebellar SAS. This agrees well with dilution of protein rich CSF from SAS before it enters the spinal canal and eventually reach the sampling site in the lumbar spine.

Our data from intrathecal contrast-enhanced MRI adds to this picture. If we assume that at 72 hours after intrathecal administration of Gadobutrol, the system is well mixed, then the concentration gradients may reflect the constant dilution of CSF in SAS with fresh CSF produced by the choroid plexus in the ventricles. At 72 hours we find the Gadobutrol concentration in cerebral SAS to be more than twice that of cerebellar SAS in good agreement with the steeper regression slopes found for proteins.

There was a large variability in these data which may be related to the large variation in CSF-dynamics among healthy controls. One volunteer showed almost no enhancement in CSF at any point after the intrathecal contrast administration. This same individual was among the highest in lumbar protein concentration while R1 in both cerebellar and cerebral SAS were among the lowest observed. This subject was removed from the analysis by outlier detection.

The large variation observed in CSF enhancement after intrathecal administration of Gadobutrol even in presumably healthy volunteers poses a challenge to the analysis of gMRI data. It is evident that the availability of Gadobutrol outside parenchyma to be studied may vary substantially between individuals. Such large interindividual variation in gadolinium enhancement has also been reported in gMRI studies [9] and suggest that the glymphatic system needs to be modelled in a subject-specific manner [9]. To enable robust quantitative analysis, this concentration must be measured individually. We argue that with the presented method such measurements may be obtained with minimal partial volume effects due to both high resolution and long echo time.

With the heavily T2-weighted data, one may segment voxels where CSF constitutes only a small volume fraction. We did not account for this in the current volume estimations, and this may have caused overestimation of the CSF-volumes On the other hand, this may allow assessment of R1 in structures smaller than the voxel size, such as perivascular spaces.

The observed ventricular reflux of CSF tracer together with the estimated protein gradients highlights the dynamic and complex nature of CSF transport. The protein gradients indicated a steady production of CSF with less protein content in the choroid plexus while the ventricular reflux of CSF tracer is probably caused by the mixing induced by cardiac and respiratory pulsations. In particular, cardiac and respiratory pulsations [34] in the aqueduct of around 1mL/s are large bidirectional flows with the potential to induce significant mixing. The unidirectional flow associated with choroid plexus production of around 0.5 L/ day or 0.006 mL/s is small in comparison, however both the bidirectional mixing and the unidirectional production may be important. In a single-subject CSF-tracer simulation study [35] (it was shown that the small choroid plexus production combined with mixing potentially speed up clearance of tracers from years to days.

To our knowledge, the relaxivity of Gadobutrol in human CSF has not been measured. Szomolanyi et al. [36] estimated relaxivity in blood plasma to be 4.97 (mM s)^−1^. It has been reported that Gadobutrol relaxivity in water is lower than in plasma [37]. Relaxivity is dependent on exchange processes and molecular thumbling rate, both which can vary depending on other solutes. Goetschi et al. [38] estimated a series of relaxivity enhancement coefficients to model the effect of plasma protein concentrations on contrast agent relaxivity. These exact concentrations may vary in CSF and we chose the more pragmatic approach of stating a relaxivity 4 (mM s)^−1^ for our analysis.

To achieve user-independent, reproducible results, we chose to use automated segmentation in this study. Available automated segmentation tools primarily focus on segmentation of the brain tissue [39] [40] [41] [42], but lack functionality in precisely segmenting CSF compartments, particularly the SAS. While Yamada et al. [43] have made progress with automated segmentation, their method is constrained to fixed SAS regions using a deep-learning approach. We chose to develop a dedicated segmentation pipeline using freely available tools in FreeSurfer. The method provides reliable delineation of CSF spaces, particularly within the SAS. This segmentation facilitates consistent and reproducible quantification of T1 relaxation times across varied compartments and is adaptable to other MRI protocols. Furthermore, the approach may easily be adjusted to make use of a segmentation with different labels than the FreeSurfer-atlases, if other SAS grouping is desired.

### LIMITATIONS

The sample size, consisting of only ten healthy controls, limits the generalizability of our findings across broader populations and specific pathological conditions. Consequently, further studies encompassing diverse cohorts and larger sample sizes are essential to validate and extend these results.

The focus on healthy individuals may not capture the range of CSF dynamics present in various neurological disorders, where anatomical and biochemical alterations could influence CSF flow differently. Additionally, while the T2W-mixed IRSE sequence and automated segmentation offer improved sensitivity and specificity, the CSF mask from the automated segmentation may include several erroneously segmented voxels. Especially along the mask’s boundary, partial volume effects or voxel spread [44] make it difficult to distinguish the exact interface between CSF and its surroundings. For the statistical analyses reported in the article, we excluded the 30% highest and lowest R1 values to ensure a robust mean R1 and eroded the CSF mask to reduce the impact of potentially mis-segmented voxels along the interface. However, this erosion may cause a bias towards data points within regions with a larger width. Furthermore, non-CSF structures with a high T2-weighted signal may be incorrectly interpreted as CSF, although most of these will be removed since we keep only the largest connected region of the CSF mask. CSF segmentations were visually inspected, but we did not conduct a quantitative study of the segmentation quality, as we consider this to be outside the scope of this article.

## CONCLUSION

Using the T2W-mixed IRSE sequence, we obtained R1 relaxation maps of cranial CSF spaces with 1 mm isotropic resolution. Intrinsic R1 of CSF in SAS was significantly correlated to lumbar protein concentrations and regression slopes mirror the mixing of CSF with high protein concentration in cerebral SAS with fresh CSF from ventricles. Spatio-temporal distribution of Gadobutrol in CSF after intrathecal administration showed a large variation among healthy volunteers. In late phase Gadobutrol concentrations mirror the CSF mixing process similar to what was seen for protein regresson.

## Data Availability

Data from the GRIP cohort are stored at Services for sensitive data (TSD) at the University of Oslo (UiO) and is publicly unavailable. An example dataset will be made available, including all MRI sequences. The code for automatic segmentation of the subarachnoid space is publicly available at: https://github.com/jorgenriseth/grip-mixed.git

## Supplementary

**Supplementary FIGURE 1.**
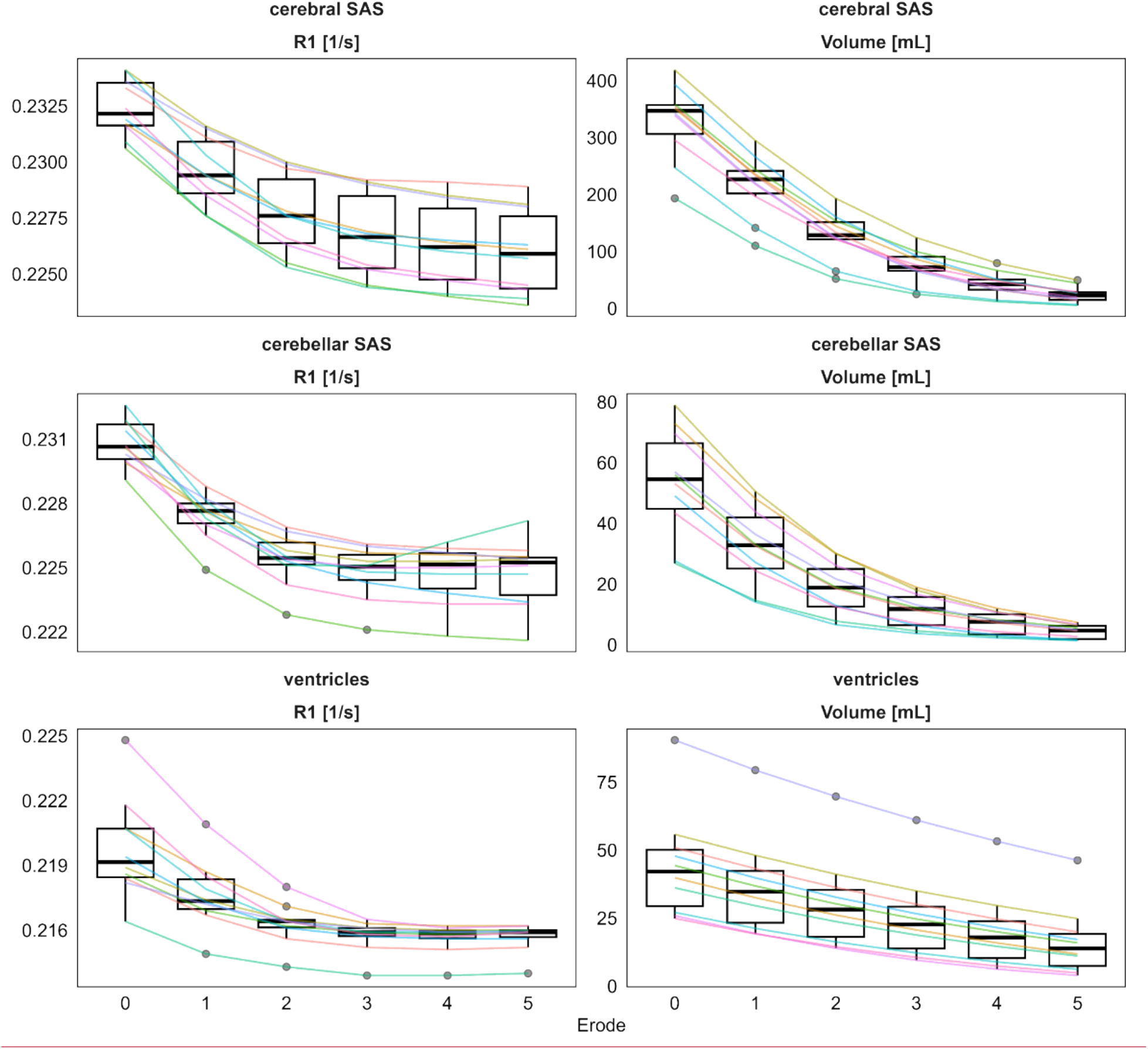
Box plots show mean R1 (left) and segmentation volume (right) after repeated erosion operations on the segmantations. In all three segmentations R1 drop as the outer voxels are removed.

